# Disparities in adherence to antiretroviral therapy of the selected subpopulations among adults living with HIV/AIDS in Tanzania. Analysis of a population-based survey 2016-17

**DOI:** 10.1101/2023.08.24.23294553

**Authors:** Dayani Adam, Ramkumar T. Balan, Abbas Ismail

## Abstract

Adherence is a critical component of successful HIV treatment among people living with HIV/AIDS. It is essential for achieving and maintaining viral suppression, which is the primary goal of HIV care and treatment. The objective of this study was to assess the disparity of the treatment adherence of selected subpopulations among HIV adult patients in Tanzania. The study used a cross-sectional survey of 792 adults living with HIV/AIDS from 31 regions in Tanzania. A two-stage stratified cluster design was used in this survey and included jackknife replicate sampling weights. Frequency tables displaying frequencies, weighted proportions, and confidence intervals were employed to depict socio-demographics by treatment adherence. Treatment adherence was calculated by subtracting missed doses from prescribed doses, dividing the result by the prescribed dosage, and multiplying by 100 to establish a percentage. Rao-Scott chi-square was used to produce the weighted proportions of the adherence within subpopulations. Also, the Adjusted Wald Test was used to examine whether or not the subpopulations are similar to or different in terms of their level of treatment adherence. The results showed that there was a significant difference in treatment adherence between the selected subpopulations. The majority of the selected subpopulations, including those based on age, sex, marital status, and level of education, were found to have disparities in the level of treatment adherence. However, the study also revealed no disparities in treatment adherence between urban and rural settings, married and widowed, age groups of 25 to 34, and those of 55 and over. The study’s findings revealed considerable disparities in treatment adherence between various subpopulations, emphasizing the necessity for specifically developed interventions to address these disparities to attain better health outcomes among adults living with HIV/AIDS.

## Introduction

The Human Immunodeficiency Virus (HIV) has been a global disease for decades since the 1980s [1,2]. That is, the disease has become a global disaster that spreads rapidly and affects a large number of people. According to [3], the disease has been reported to have infected 79.3 million people worldwide and 36.3 million people have been killed since its outbreak. The most recent report shows that there are 650,000 deaths caused by Acquired Immune Deficiency Syndrome (AIDS) by the end of 2021, with 1.5 million new HIV infections[4]. About 36.7 million (95.5%) people living with HIV (PLHIV) are adults, while 1.7 million (4.5%) are young children with an age range between 0 to 14 years [4,5].

Due to the coverage, effects, and reoccurrence of new infections of HIV/AIDS, The Joint United Nations Programme on HIV/AIDS (UNAIDS) and other related stakeholders in combating the disease prioritize treatment programs. That is, the diagnosis of new HIV infections necessitates the intensification of the use of antiretroviral therapy (ART) for the good health and well-being of the PLHIV being one of the sustainable development goals strategies to end AIDS by 2030 [5]. This is in line with the adherence to ART as the way HIV patients follow and respect the medical advice given to them at the right time, dose, and manner[6].

Adherence to ART has been an essential strategy for clinical success especially viral suppression among people with HIV/AIDS. Achieve a good virological and immunological status, it requires adherence to be above the recommended level from the UNAIDS (>95%) [7]. This results in viral load suppressions and minimizes new infections. Nevertheless, the initiation of ART aims to boost cluster differentiation 4 (CD4) cell counts and reduce viral loads that lead to lowering PLHIV morbidity and mortality[8–10]. The President’s Emergency Plan for AIDS Relief (PEPFAR) has numerous funding and support for the ART program in sub-Saharan countries even though adherence to ART, viral loads, and drop in new infection levels are unsatisfactory for the PLHIV[11]. According to [12], the proportion of PLHIV in Nigeria experiencing poor adherence is on the rise, and that resulted from most of the PLHIV experiencing immunological and virological failure. Also, the study by [13] in Botswana reported that there is a positive correlation between good adherence, CD4 increase, and viral load suppressions.

Over time, Tanzania’s access to ART therapy and care has scaled up. This has led to improvements in adherence and clinical outcomes across all age groups[14]. The report from [15] asserts that 52.2% of HIV adult patients aged 15 to 64 know their status, and 93.6% of them are receiving ART and experienced viral suppression by 87.0%. The advent of ARVs in 2003 has significantly caused improvement in ART tactics and activities. The study by [16] reported that forgetfulness, young age, and perception of well-being, limit ART adherence which sometimes impedes therapeutic services in Tanzania.

Studies have reported that disparities in treatment adherence exist among various subpopulations. Most of the disparities were identified in sex, age, marital status, employment status, and residential settings. For instance, studies by [17,18] showed that females were less adherence than males receiving ART indicating that there is a disparity in treatment adherence among the sex of the adult patients. Also the study by [19] that participants with older ages have higher adherence compared to those with low age. The disparity is commonly also found in the residential settings where identified that the patients who lived in urban areas were more adhered compared to the patients who lived in rural areas [20,21]. The other study revealed that married patients were identified to be more adhered compared to those with single and separated [22]. Additionally, disparities in treatment adherence may vary across different medical conditions and treatments. Knowing disparities may help to improve intervention. This study aims to determine and assess the disparity of treatment adherence prevalences of the ART of the selected subpopulations among adults living with HIV/AIDS.

## Materials and Methods

### Study design, Setting, and Population

This study analyzed cross-sectional data from Tanzania HIV Impact Survey (THIS) collected from 31 regions of the United Republic of Tanzania in 2016-2017 for both Tanzania Mainland and Zanzibar. The Population-based HIV Assessment (PHIA) measures HIV prevalence and incidence with a high HIV burden and is funded by the President’s Emergency Plan for AIDS Relief (PEPFAR) through the US Centers for Disease Control and Prevention (CDC). The population consisted of HIV adult patients aged 15 and above on ART care.

### Sources of data

The study applied the secondary data from THIS national representative survey to provide estimates of HIV risk, burden, and efficiency of ARV in the treatment process obtained from both biomarker and individual datasets. These are the national representative surveys that are designed to collect data on the key biological endpoints to provide estimates of HIV risk, burden, and efficiency of ARVs in the treatment process. PHIA-related deterrence, care, and treatment interventions were executed in the country by using a questionnaire administered for both biomarkers and individuals including HIV and syphilis. As the extracted datasets were identified as two distinct datasets, namely the individual and biomarker datasets, they were combined to create the single dataset used for analysis in this study.

### Sample, Sample Design, and Sampling Weights

The sample size for this study was 792 HIV adult patients who were on ART care extracted from 34,060 participants from both biomarker and interview datasets. A two-stage stratified cluster design was used for the sampling procedure. This design was taken from a PHIA survey of 31 regions that are viewed as strata. Enumeration areas that make up the initial step of sampling total roughly 526 and were divided into clusters as primary sampling units. Primary sample units were designated as the first stage and were selected using probabilities proportionate to the number of homes based on the denominator of the 2012 population census. The second stage involved a random systematic sample taken at rates to yield-self weighting was used in the second step of sampling to choose households from the first stage of sampling.

Due to this, the study integrated sampling weights known as jackknife and replicates weights found in the datasets to get unbiased results. The jackknife replicate weights variables should be reviewed to see whether they should be included in the relevant analysis for each study, depending on whether they came from the either interview or biomarker dataset. Each dataset has the initial jackknife replicate weight and 257 replicate weights. These weights ensure that the analysis appropriately represents the target population and produces accurate population-level results by accounting for the different probabilities of selection at each stage of sampling. Hence, by accounting for the complex sampling design and predicting sampling errors, jackknife replicate weights are used to get unbiased results. The details on the sampling were provided in the PHIA data use manual [23].

### Study Variables

Treatment adherence was an outcome variable, calculated based on how frequently patients missed doses in the previous 30 days. This assessment gave an indicator of the extent to which patients followed their treatment regimen over the duration they were given. The calculation of adherence to ART is detailed in equation (1).

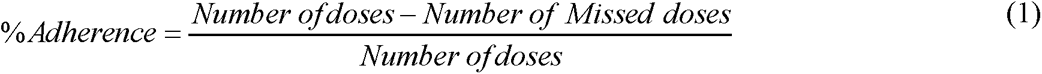

The treatment adherence was then classified as adhered or not adhered depending on whether it was above or below 95%. The subpopulations based on sex, age, marital status, education level, residential settings, and employment status were employed to create the matched pairs of adherence to test for disparity.

### Data extraction

The 2016-17 THIS household dataset was downloaded in Stata format with permission from the PHIA project website. After understanding the detailed datasets and coding, further data recording was carried out to modify variables, clean the data, and align it with the study. The data recording process aimed to ensure accuracy and reliability, enhancing the dataset’s suitability for analysis and interpretation. We obtained the PHIA datasets from the PHIA Project website at https://phia-data.icap.columbia.edu/datasets?country_id=10.

### Data Analysis

Stata version 17 and Microsoft Excel were used in the analysis of the survey. The complex survey’s data were declared using the **“svyset”** command. This enables integration of the two-stage stratified cluster design features in the Stata software to allow data to be analyzed as survey data. The incorporation of the “**svy**” utilized a complex survey design in each of the analyses done in the study. Using “**test**” as a post-estimation command, the disparity in treatment adherence rates among particular subpopulations was examined. The descriptive analysis procedures employed frequencies, tables, percentages, graphs, means, and proportions. Frequency tables displaying frequencies, weighted proportions, and confidence intervals were employed to depict participant characteristics concerning their HIV status. Additionally, a bar graph was employed to display the adherence status of HIV adult patients. The study also employed an adjusted Wald Test and Rao-Scott chi-square. Rao-Scott chi-square was used to determine the association between the level of adherence and socio-demographic characteristics. Adjusted Wald statistical test was used to examine whether or not the matched pairs of subpopulations were similar to or different in terms of their level of treatment adherence. Adjusted Wald Test was employed to test the overall equality adherence, as well as matched pairs of sex, age, marital status, residential settings, employment status, and level of education by HIV adult patients.

### Ethical consideration

Following receiving the requisite permission from the PHIA Data Manager, anonymized open-access website data for this study was collected from the PHIA website. By refraining from accessing personally identifiable information throughout the data extraction process, such as file numbers, the researcher was able to guarantee the privacy and confidentiality of the data. A thorough assessment and approval of all PHIA survey protocols, including permission forms, screening forms, refusal forms, referral forms, recruitment materials, and questionnaires, was required for the data collection process. The study protocol underwent rigorous review and approval by the institutional review boards (IRBs) of Columbia University Medical Center, Westat, CDC, the National Institute for Medical Research, and the Zanzibar Medical Research and Ethics Committee. These IRBs played a vital role in ensuring the ethical conduct of the study and protecting the rights and well-being of the participants involved.

## Results

### Baseline characteristics of the participants of the survey

The survey included a total of 34,060 participants, consisting of individuals aged 15 or older. Gender distribution among the participants revealed that 51.2% identified as female, while 48.8% identified as male. HIV status evaluation was conducted for a subset of participants, with 90.3% (30,740 individuals) testing HIV-negative, indicating the absence of the virus. However, 5.6% (1,895 individuals) tested HIV positive, indicating that they were infected with the virus whereas the rest they don’t know their refused to have the test for HIV. Among those who tested positive for HIV, 41.8% (792 individuals) were receiving ART, a treatment method for managing HIV infection. These baseline characteristics provide valuable insights into the demographics and HIV status of the survey participants at the time the study was conducted.

### Prevalence of Treatment Adherence to Antiretroviral Therapy among HIV adult patients

The proportion of days missed dose in the previous 30 days of treatment was used to calculate the patient’s level of adherence. The data revealed that 612 (77.3%) patients had full adherence, 79 (9.9%) had one day missed dose, 57 (7.2%) had two days missed doses, and 44 (5.6%) had three days missed doses in the previous 30 days of the treatment. As the total adherence of 792 PLHIV getting ARVs, the mean adherence in this survey was 98.7% (95% CI; 98.5-98.9). The 95% adherence rate, as recommended by the WHO was used to dichotomize adherence. The terms “patient adhered” and “patient not adhered” relate to optimal and suboptimal adherence respectively. The optimal adherence level is defined as greater than or equal to 95%. Furthermore, among HIV male patients receiving ART treatment, 182 (83.9%) of them took their medications as prescribed compared to 35 (16.1%) of them who did not. For HIV female patients receiving ART treatment, 509 (88.5%) of them took their medicine as directed compared to 66 (11.5%) who did not. Figure 1 provides an information summary.

**Figure 1.**
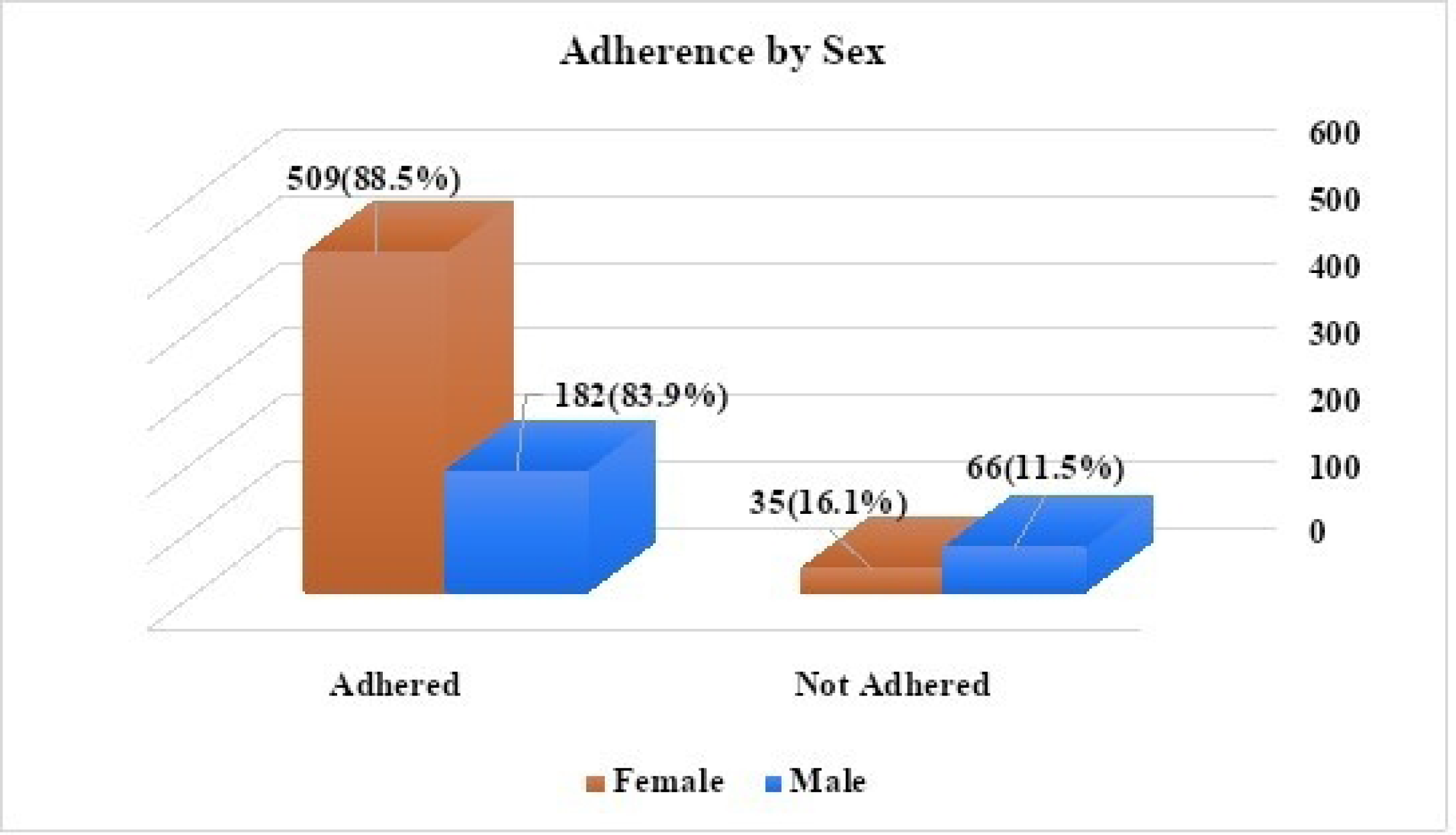
Adherence by sex.

### Prevalence of ART treatment adherence across levels of socio-demographic factors

The results from Rao-Scott chi-square statistical test found that there was no significant association between the level of adherence and selected socio-demographic variables in the bivariate analysis. Also, the aforementioned statistics produced weighted proportions of adherence among all socio-demographic subpopulations. The results show that It has been indicated that male and female adherence rates were 86.4% (95% CI: 79.5-91.5) and 88.9% (95% CI: 85.2-91.8) respectively. The highest prevalence of adherence 91.0% (95% CI: 85.8-94.5) was seen among patients between the ages of 45 and 54. The patients who got secondary education and above had a prevalence rate of 88.6% (95% CI; 84.9-91.5). But, those with no education or training at all had 83.3% (95% CI; 68.8-91.9), and the patients with no education and pre-primary education had the highest prevalence. Patients who were either divorced or separated had adherence prevalences of 90.4 (95% CI: 83.8-94.5) and 90.3 (95% CI: 76.8-96.4) respectively. In rural settings, the prevalence was 87.0% (95% CI: 81.8-90.9). This rate runs contrary to the town where it was 89.3% (95% CI: 85.3-92.3). Employed patients had higher adherence rates than unemployed ones. The prevalence of treatment adherence to ART on socio-demographic factors is summarized in Table 1.

**Table 1:**
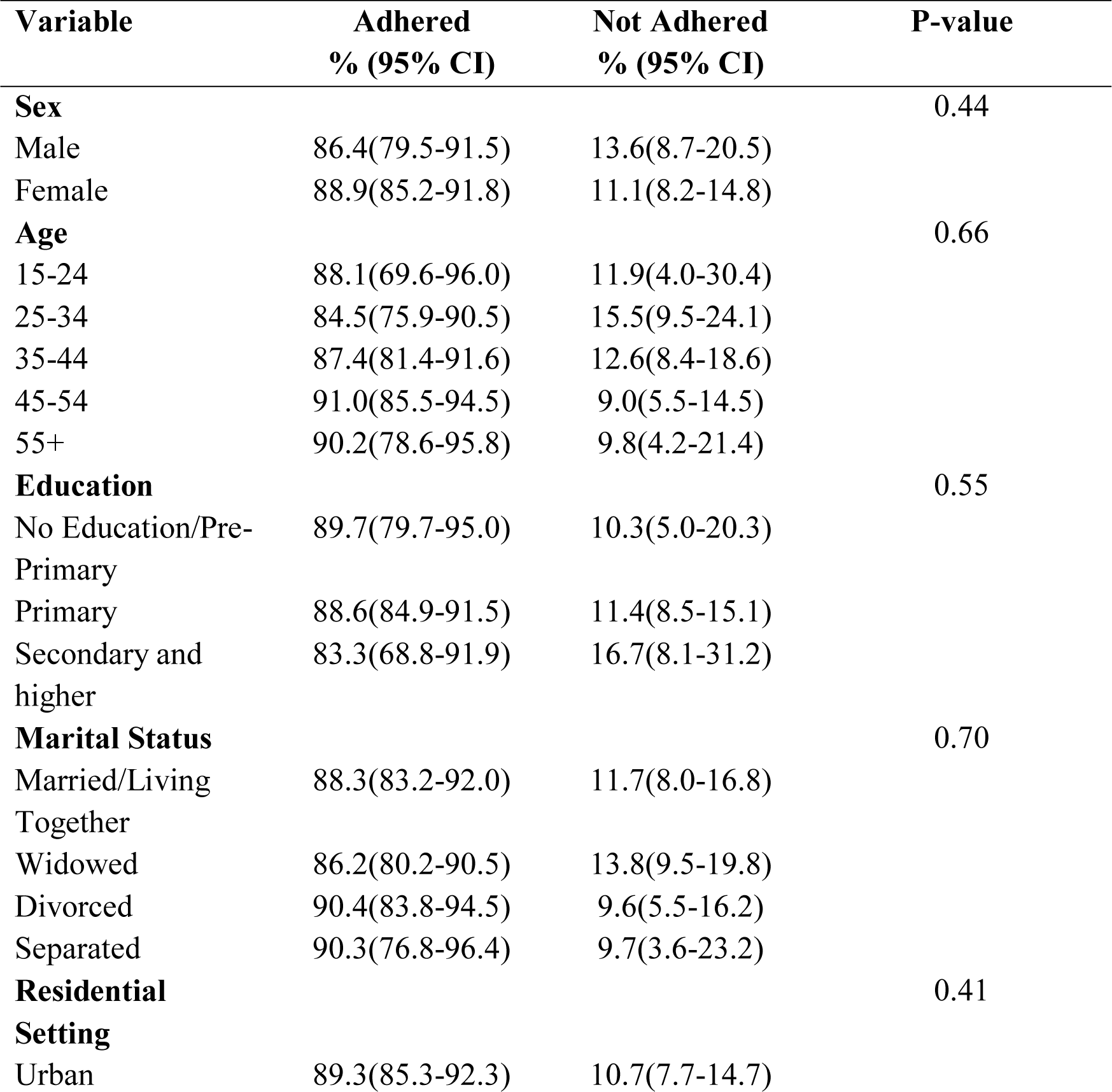
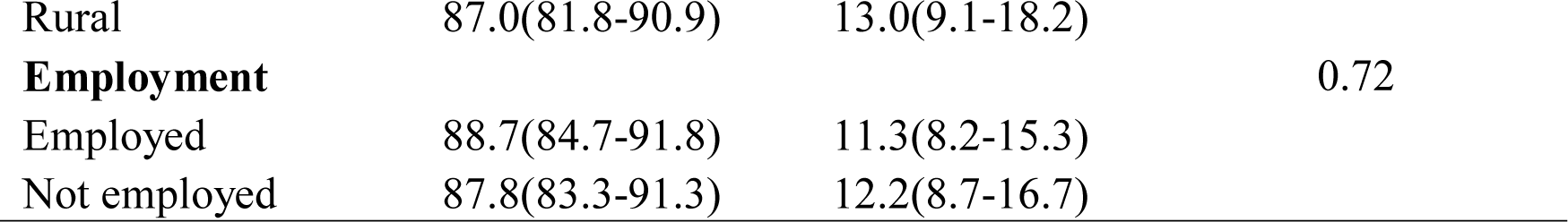
Prevalence of Treatment Adherence to ART on Socio-Demographic Variables.

### Disparity of treatment adherence to antiretroviral therapy

Primarily, the overall equality of prevalence of adherence with the hypothesized value of 0.95 is recommended by UNAIDS. The results showed that treatment adherence was 98.7% indicating that it was statistically higher than 0.95 (F = 22.32, p < 0.001). This indicates that the level of adherence calculated achieved its optimal adherence.

Also, the matched pair of the sex subpopulation was formulated to test the disparity of the prevalence of treatment adherence between females and males. The results showed that patients’ adherence was significantly different from female and male patients (F=99.15, p<0.001). This shows that there is a disparity in treatment adherence between male and female patients. On the other hand, the matched pairs of the age categories were formulated and tested the disparity among different age subpopulations. The results showed that on the matched pairs constructed, three results were shown. Between various age groups, the study discovered significant statistical disparities. Ages 15–24 years differed considerably from 25–34, 35–44, 45–54, and 55 and above years. When compared to the 35-44 age group, the same tendency was seen for the 25-34 age group. The 35-44 age group was very different from the 45-54 and 55 and above age groups. The 45-54 group, in addition, differed significantly from the 55 and above years’ group. The statistical studies provided significant support for these differences, typically with p-values below 0.0001, demonstrating that different age groups exhibit distinctive behaviors on treatment adherence. Only patients in the age group of 25-34 were revealed to have the same prevalence of adherence as the age group of 55 and older (F=1.06, p=0.305).

The subpopulation based on the residential settings was constructed and tested to reveal if HIV adults receiving ART care adhered to the treatment regimen equally between rural and urban. The results showed that the prevalence of treatment adherence for ART care did not statistically differ from rural to urban settings (F=0.28, p=0.59). The study examined adherence based on marital status and found significant statistical differences between different pairs. Patients who were married showed significant differences from those who were divorced, separated, and widowed. Similarly, patients with separated marital status were significantly different from divorced and widowed patients. Widowed and separated patients, as well as divorced and separated patients, also showed significant differences from each other. The p-values were all below 0.0001, indicating strong evidence for these observed statistical distinctions in adherence based on marital status. The disparity of treatment adherence was not found between married and widowed (F=3.42, p=0.065).

The study found that patients with various levels of education showed significantly variable degrees of adherence. Patients who had pre-primary education were significantly different from those who had completed primary education (p<0.0001) and from those who had completed secondary education (p<0.05). Additionally, as compared to patients with secondary education, patients with primary education displayed significant differences in adherence (p<0.0001). This shows that education levels influence patient adherence behaviors.

Correspondingly, the matched pair of employment status subpopulations of the HIV adult patients on ART care has also been tested for their equality between those patients who are employed and unemployed. The study found that the HIV adult patients have significantly different from those employed (F=12.67, p<0.0001). Thus, the data on the disparity of treatment adherence by subpopulations are summarized in Table 2.

**Table 2:**
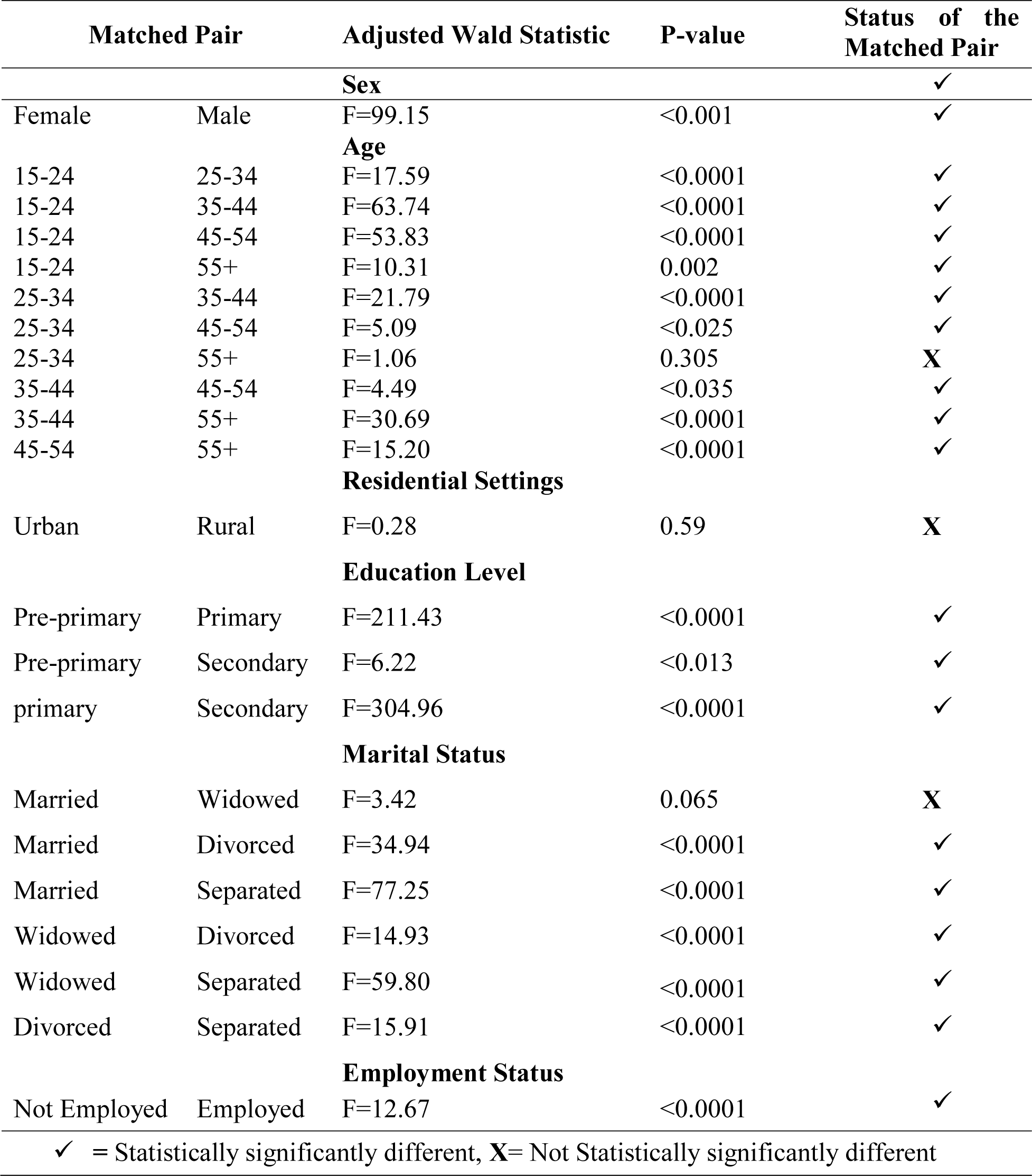
Disparity of treatment adherence among selected subpopulations.

## Discussions

The current study found a high prevalence of adherence to ART at 98.7%, which is in line with the recommended rate by UNAIDS. To successfully manage HIV/AIDS and guarantee the best possible treatment outcomes, achieving and maintaining such a high adherence rate is crucial. This result is consistent with previous studies conducted in Senegal among ART adults [24–26], which also reported a high rate of adherence. However, this result contradicts other studies by [13,27], which reported low adherence rates. These findings suggest that although the desired level of adherence has been achieved in this study, there may be variations in adherence rates depending on the population being studied and the measures used to assess adherence.

Research on disparities in treatment adherence among adults living with HIV/AIDS has been conducted extensively over the past few decades. Many studies have found that there are differences in treatment adherence between different subpopulations. ART adherence among these different subpopulations affected by HIV/AIDS has been found to vary widely in research. These subpopulations may differ in terms of a variety of demographic characteristics, including sex, age, education level, employment status, marital status, and residential settings. The ability to customize interventions and support systems to increase adherence rates among particular groups depends on an understanding of these disparities.

The current study found that the overall prevalence of treatment adherence was statistically higher than 0.95, which is in agreement with a previous study by [28] that reported a similarly high rate of adherence. However, this study contradicts a study by [21] that reported a lower rate of adherence than the recommended rate by UNAIDS. These findings suggest that while the overall prevalence of adherence may be high, there may still be variations in adherence rates among different subpopulations

In the present study, significant disparities in treatment adherence across male and female subpopulations were found, with female adherence being significantly from male. This result is in line with studies by [17,18,29] that found females and males have disparities in their level of adherence. The studies by [30–32], in contrast, found no statistically significant differences between male and female adherence rates. This shows that disparities in adherence between men and women may be the cause of gender differences in adherence.

The results of the current study indicate that there is a disparity in terms of age in treatment adherence because the majority of age-matched subgroups showed statistically significant differences in adherence rates. This result is similar to a cross-sectional study by [19,33], which also found significant differences in adherence rates among age subgroups. The studies by [32,34] that did not discover any significant variations in adherence rates between older and younger persons contradict this finding. These inconsistencies could be explained by variations in the way age groupings were created and characterized in the research. Generally, these data imply that age may influence treatment adherence and that it should be taken into account in programs designed to increase adherence. The study found significant disparities in treatment adherence between HIV/AIDS patients who were employed and those who were not. This is consistent with a study [36], which found no discernible change based on occupation. Studies [31,35] on the other hand reported on adherence disparities among employed and unemployed HIV adult patients. These distinctions could result from various interpretations and evaluations of employment. However, these results highlight the need of taking job status into account as a possible driver of treatment adherence in focused treatments.

The study shows that there are disparities in adherence to ART treatment based on educational attainment. Patients’ adherence patterns varied between those with pre-primary education and those with primary education, and between those with primary education and those with secondary education. This implies that different levels of education influence treatment strategies and adherence rates within ART programs. The study by [37], aligns with the results from the current study. These findings are inconsistent with the studies by [31,32,38] who indicated that the level of education does not statistically differ in terms of adherence between different groups. This suggests that various aspects of differentiation have contributed to these differences.

Based on the residential setting subpopulation, the findings from the current study revealed that the treatment adherence among the two areas was statistically insignificant different. This research supported the [31,39–41] studies, which found that there were no significant differences in adherence rates between rural and urban residents. This indicates that there is no disparity in treatment adherence among HIV adult patients living in urban and rural areas. These findings are contrary to the findings of [20,21,42], who reported that there were disparities in adherence in urban and rural settings. The results of this study revealed that treatment adherence varies significantly among matched pairs of different marital statuses, except for married and widowed pairs. This result aligns with [22,43–45], which showed that married HIV adult patients had a significant disparity in treatment adherence compared to unmarried and widowed HIV adult patients. This finding was contrary to the study by [46] in Ethiopia asserted that only married appeared to significantly have disparity in adherence and widowed. In the context of HIV/AIDS, the study discovered disparities in treatment adherence rates among various subpopulations, underscoring the significance of monitoring adherence and figuring out potential causes of poor adherence rates in particular populations. Therefore, it is important to continue monitoring adherence to ART and to identify and address factors that may contribute to low adherence rates in certain populations.

## Conclusions

This study has demonstrated the differences in treatment adherence across several subpopulations, including age, sex, marital status, education level, and work position. To attain optimal adherence among the identified groupings, these findings provide information that can assist the ministry and other stakeholders in eliminating and improving low ART adherence among subgroups to achieve viral load suppression among patients living with HIV/AIDS. This makes it possible for other researchers to identify the reasons behind variances in the affected subpopulations.

## Data availability

Data from the Tanzania HIV Impact Survey were utilized in this study. The data can be retrieved from https://phia-data.icap.columbia.edu/datasets as the PHIA website to request these data.

## Acknowledgments

The authors would like to thank THIS for allowing access to the dataset,https://phia-data.icap.columbia.edu/datasets.

